# Impact of intraoperative use of venovenous extracorporeal membrane oxygenation on the status of von Willebrand factor large multimers during single lung transplantation

**DOI:** 10.1101/2023.02.07.23285614

**Authors:** Hisashi Oishi, Yoshinori Okada, Yamato Suzuki, Takashi Hirama, Yutaka Ejima, Shin-ichi Fujimaki, Shingo Sugawara, Noriyuki Okubo, Hisanori Horiuchi

## Abstract

**Purpose:** von Willebrand factors (vWFs), hemostatic factors, are produced as large multimers and are shear stress-dependently cleaved to become the appropriate size. A reduction in vWF large multimers develops in various conditions including the use of extracorporeal life support, which can cause excessive-high shear stress in the blood flow and result in hemostatic disorders. The objective of this prospective study was to investigate the impact of venovenous extracorporeal membrane oxygenation (VV ECMO) use on the status of vWF large multimers and hemostatic disorders during single lung transplantation (SLT).

**Methods:** We prospectively enrolled 12 patients who underwent SLT at our center. Among them, seven patients were supported by VV ECMO intraoperatively (ECMO group) and the remaining five patients underwent SLT without ECMO support (control group). The vWF large multimer index (%) was defined as the ratio of the large multimer proportion in total vWF (vWF large multimer ratio) derived from a patient’s plasma to that from the standard human plasma.

**Results:** The vWF large multimer index at the end of the surgery was significantly lower in the ECMO group than in the control group (112.6% vs. 75.8%, respectively; *p* < 0.05). The intraoperative blood loss and the amounts of intraoperative transfusion products in the ECMO group tended to be greater than those in the control group; however, the differences were not significant.

**Conclusion:** During SLT, the intraoperative use of VV ECMO caused a decrease in the vWF large multimer index. However, the vWF large multimer index was maintained at > 75% in average at the end of SLT, which did not affect the bleeding complications.

## Introduction

One of the most severe complications in lung transplantation is perioperative blood loss. Large-volume transfusions of red blood cells (RBCs) and platelets have been reported to be associated with various complications in lung transplant recipients in multiple studies [1]. Diamond et al. have identified large-volume RBC transfusion (> 1 L) as a risk factor for grade 3 primary graft dysfunction (PGD) in a prospective, multicenter cohort study [2]. Platelet transfusions were shown to be substantial risk factors for the production of anti-human leukocyte antigen (HLA) antibodies [3], which have a potential to cause chronic lung allograft dysfunction (CLAD)[4].

Intraoperative use of extracorporeal life support (ECLS) is one of the risk factors for the increased amount of bleeding and transfusion in lung transplantation. Among ECLSs, cardiopulmonary bypass (CPB) had been reported to increase blood transfusion due to coagulopathy and inflammatory response [5]. Currently, venoarterial extracorporeal membrane oxygenation (VA ECMO), which is associated with lower blood loss during lung transplant surgeries compared with CPB [6], is preferably used for lung transplant cases unless intracardiac repairs are required [7]. Moreover, venovenous (VV) ECMO has been used as an intraoperative extracorporeal mechanical support for lung transplantation at some lung transplant centers in recent years [8]. We started utilizing VV ECMO in 2015 and reported that VV ECMO could be a choice for intraoperative extracorporeal mechanical support during single lung transplantation (SLT) [9].

Although the increase in the amount of bleeding during ECLS is primarily due to the use of heparin, recent studies have suggested the involvement of acquired von Willebrand syndrome (AvWS), which is one of the hemostatic disorders. von Willebrand factor (vWF) is a hemostatic factor that is produced and secreted as large multimers by endothelial cells and megakaryocytes. vWF initially forms a huge multimer and is then shear stress-dependently cleaved in the bloodstream by its specific cleaving enzyme ADAMTS13, and then is present as multimers consisting of 2 – 80 vWF monomers [10,11]. The conformation change of A2 domain of vWF by shear stress in the blood flow triggers its cleavage by ADAMTS13. Among the vWF multimers, higher molecular weight multimers (large multimers) are known to play a critical role in hemostasis [10,11]. Lack of ADAMTS13 results in excessive platelet thrombus formation by the function of ultra-large vWF multimers, as seen in patients with hereditary thrombotic thrombocytopenic purpura [12]. On the other hand, some genetic mutations in the vWF A2 domain cause a reduction in large multimers and induce a hemostatic disorder by increasing the susceptibility of vWF to cleavage by ADAMTS13 [11]. A reduction in vWF large multimers develops in various other conditions including cardiovascular diseases such as aortic stenosis and the use of ECLS, which can cause excessive-high shear stress in the blood flow and result in hemostatic disorders [11]. This hemostatic disorder caused by excessive cleavage of vWF is defined as AvWS [13].

Kalbhenn et al. reported that patients treated with VV ECMO support developed AvWS [14]. They have shown that using VV ECMO for pneumonia, ARDS, and so forth can cause unphysiologically high shear stress in the blood stream leading to AvWS within 24 hours after ECMO exposure [14]. However, it remains unclear whether VV ECMO use in a short period of time during lung transplant surgery can causes a decrease of vWF large multimers. The objective of this prospective study was to investigate the impact of VV ECMO use on the status of vWF large multimers and hemostatic disorders during SLT.

## Patients and Methods

### Patients, blood sample and data collection, and study groups

The Ethics Committee of Tohoku University Graduate School of Medicine approved this retrospective study (IRB approval number, 2016-1-629; protocol number, UMIN000018135), and written informed consent was obtained from all participants. We prospectively enrolled 12 patients who would undergo SLT under VV ECMO or without ECMO at Tohoku University Hospital from April 2017 to December 2019 (participant recruitment start date, January 10, 2017; last follow-up date, December 31, 2020) (Fig. 1). The study was carried out in compliance with the 2000 Declaration of Helsinki and its later amendments.

**Figure 1.**
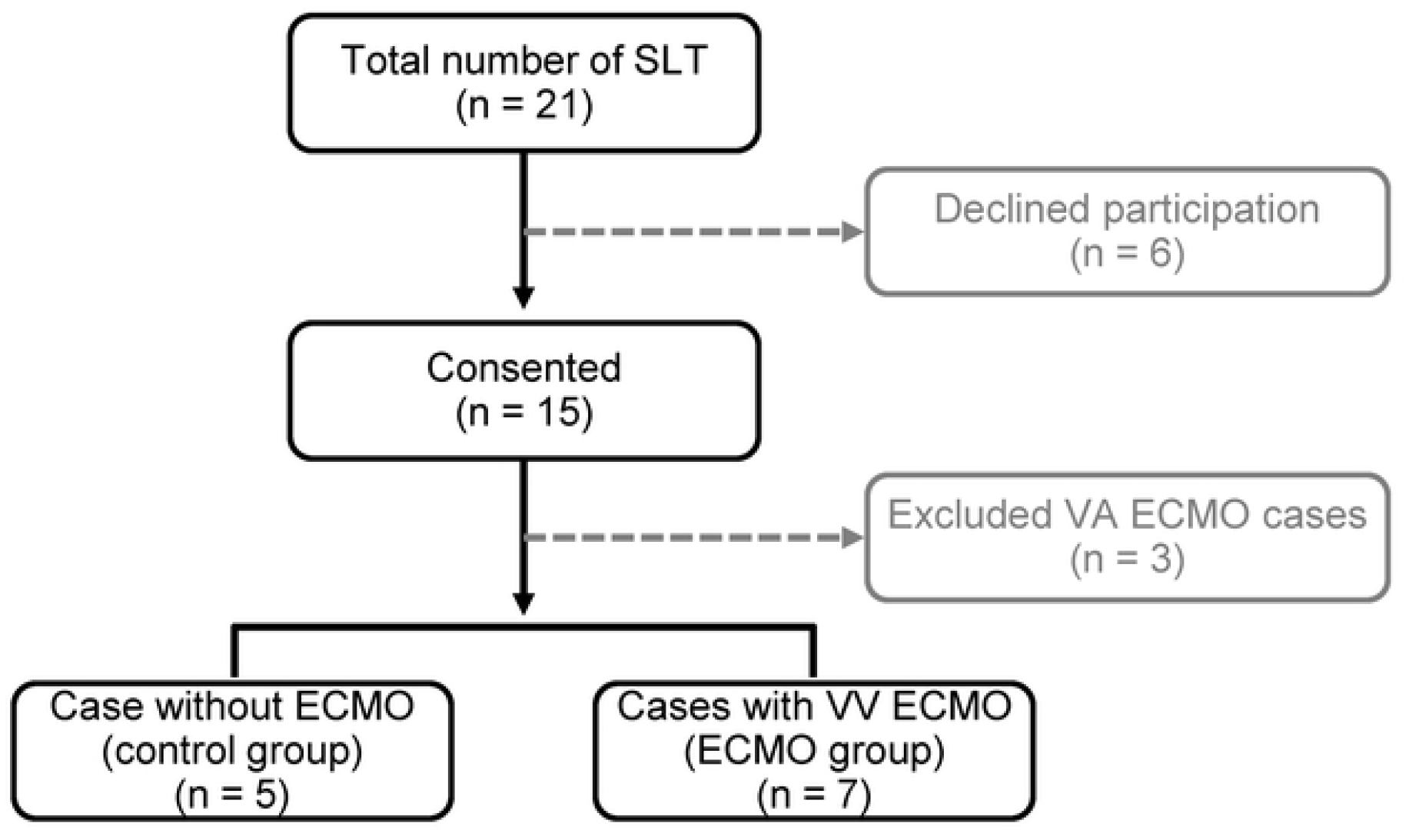
Flow diagram for study participation. SLT, single lung transplantation; VA ECMO, venoarterial extracorporeal membrane oxygenation; VV ECMO, venovenous extracorporeal membrane oxygenation.

For the analysis of ADAMTS13 activity and vWF large multimers, blood samples were collected in a tube containing 3.2% sodium citrate from the patients immediately before lung transplantation, before reperfusion (immediately before reperfusion of the lung graft), and at the end of surgery. The plasma of the collected blood was stored at - 80 °C until analysis. The collected data were as follows: pretransplant demographics of the recipients, operative characteristics, postoperative characteristics, and mortality. The level of the pleural adhesion was assessed intraoperatively and graded as follows according to the area of adhesion: severe, 50% to whole thoracic cavity; moderate, partial (only band adhesions) to < 50%; mild, no to partial adhesion; none, no adhesion.

### ECMO induction procedure and intraoperative management

We established VV ECMO by using right and left femoral veins for the cannulation sites. For the inflow, a 15–22 Fr cannula (CAPIOX EBS, Terumo Corp., Tokyo, Japan or HLS Cannulae, Getinge Group Japan K.K., Tokyo, Japan) was inserted, and the tip was adjusted within a caudal part of the inferior vena cava to minimize the recirculation. For the outflow, a 15– 21 Fr cannula (CAPIOX EBS, Terumo Corp., Tokyo, Japan or HLS Cannulae, Bio-Medicus, Medtronic Japan Co., Ltd., Tokyo, Japan) was inserted and the tip was placed at the right atrium using transesophageal echocardiography guidance. The blood flow rate of VV ECMO was started at 50% of the estimated cardiac output of the recipient and adjusted according to the intraoperative results of the arterial blood gas analysis (1.5 – 2.5 L/min). Heparin administration was controlled to target the activated clotting time (ACT) from 160 – 180 s.

### Measurement of ADAMTS13 activity and quantitative method for calculation of the vWF large multimer index

ADAMTS13 activity was measured based on the FRETS-VWF73 assay by SRL Co. (Tokyo, Japan) [15]. We first determined the concentrations of vWF antigen (vWF:Ag) in the standard human plasma (Siemens Healthcare Diagnostics Products GmbH, Marburg, Germany) by quantitative immunoelectrophoresis using an automatic analyzer. Then, the same amount of vWF:Ag in the plasma of each sample and that in the standard human plasma were analyzed side-by-side by the so-called vWF multimer analysis, which is a western blot analysis under non-reduced conditions. The bands of the 11th and those over the 11th were defined as large multimers. The vWF large multimer proportion was calculated as the densitometric area of those bands using Image J (National Institutes of Health, Bethesda, MD). Then, we determined the parameter, “vWF large multimer index,” as described previously [16]. Briefly, the vWF large multimer index (%) was defined as the ratio of the large multimer proportion in the total vWF (vWF large multimer ratio) derived from a sample to that from the standard human plasma in the adjacent lane [16,17] (Fig. 2). Thus, a patient’s vWF large multimer index was expressed as a ratio (percentage) of the standard.

**Figure 2.**
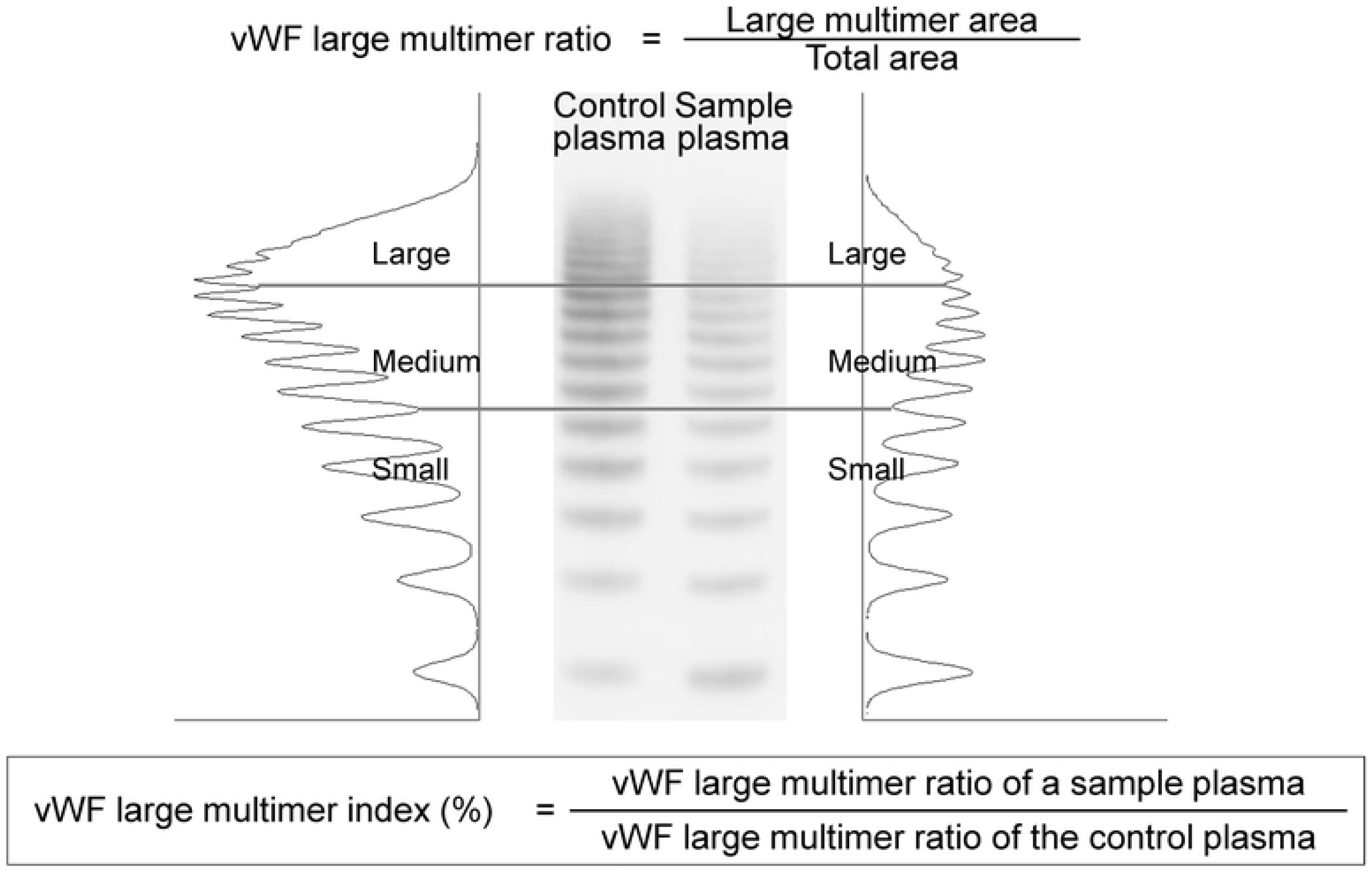
Quantification method for calculation of the von Willebrand factor (vWF) large multimer index. We first determined the concentrations of von Willebrand factor (vWF) in the standard human plasma (Siemens Healthcare Diagnostics Products GmbH, Marburg, Germany) and in each sample simultaneously by western blot analysis under reduced conditions followed by densitometry analysis. Bands 1–5 (starting from the lowest molecular weight band) were classified as low, 6–10 as medium, and those >10 as large multimers. The vWF large multimer proportion was calculated as the densitometric area of those bands. The vWF large multimer index (%) was defined as the ratio of the large multimer proportion in the total vWF (vWF large multimer ratio) derived from a sample to that from the standard human plasma in the adjacent lane.

### Statistical Analysis

Continuous data are expressed as group mean ± standard error with ranges. Categorical variables are presented as numbers and percentages. For comparison between the groups, Student’s or Welch’s *t*-test was used for the parametric data. Mann-Whitney *U* test was used for the nonparametric data. Fisher’s exact test or the chi-square test was performed for the categorical values. The ADAMTS13 activity, vWF:Ag, and vWF large multimer index at each time point were analyzed by repeated measures two-way ANOVA. Differences were considered significant at the p < 0.05 level. Statistical analyses were performed using Prism 5 (GraphPad Software Inc, La Jolla, Calif) and SPSS Statistics 21.0 for Windows (IBM Corporation, Chicago, IL, USA).

## Results

### Study groups and pre-transplant demographics of the recipients

All enrolled patients were followed up until one year after lung transplantation at Tohoku University Hospital, except for one 90-day mortality case. In the present study, we analyzed 5 SLT cases without ECMO use and 7 SLT cases with VV ECMO use. Table 1 shows the pretransplant demographics of the recipients. Recipient age, gender, height, weight, and body mass index (BMI) were similar between the 2 groups. Forced vital capacity (FVC) was comparable between the groups. Because the control group included 3 cases of chronic obstructive pulmonary (COPD), forced expiratory volume in one second (FEV1) in the control group tended to be lower than that in the ECMO group; however, the difference was not statistically significant. There was no significant difference between the 2 groups in prothrombin time–international normalized ratio (PT-INR), activated partial thromboplastin time (APTT), or platelet count.

**Table 1.** Pretransplant demographics.

|  | Control group<br>(*n* = 5) | ECMO <sup>a</sup> group<br>(*n* = 7) | *P* value |
| --- | --- | --- | --- |
| Age (years) | 48.4 ± 3.8 (34 – 57) | 47.1 ± 4.1 (23 – 59) | 0.85 |
| Gender (M/F) | 3 / 2 | 4 / 3 | 0.92 |
| Height (cm) | 164.4 ± 3.2 (153.5 – 175.0) | 162.4 ± 2.6 (150.4 – 173.0) | 0.66 |
| Weight (kg) | 50.5 ± 6.4 (35.0 – 75.0) | 55.6 ± 6.9 (30.0 – 91.8) | 0.64 |
| BMI (kg/m<sup>2</sup>) <sup>b</sup> | 18.5 ± 2.0 (13.0 – 26.4) | 20.7 ± 2.0 (13.3 – 30.7) | 0.50 |
| Indication |  |  |  |
| IPF <sup>c</sup> | 0 (0%) | 4 (57.1%) |  |
| COPD <sup>d</sup> | 3 (60.0%) | 0 (0%) |  |
| LAM <sup>e</sup> | 1 (20.0%) | 2 (28.6%) |  |
| CTD-ILD <sup>f</sup> | 1 (20.0%) | 1 (14.3%) |  |
| Pulmonary function test |  |  |  |
| FVC <sup>g</sup> (% predicted) | 55.1 ± 5.0 (46.4 – 76.0) | 50.9 ± 8.0 (23.6 – 85.0) | 0.71 |
| FEV1 <sup>h</sup> (% predicted) | 35.1 ± 7.5 (17.8 – 57.6) | 43.7 ± 6.5 (26.3 – 78.4) | 0.44 |
| PT-INR <sup>i</sup> | 0.99 ± 0.02 (0.93 – 1.04) | 1.00 ± 0.02 (0.99 – 1.00) | 0.79 |
| APTT <sup>j</sup> (sec) | 32.2 ± 2.7 (27.4 – 33.9) | 30.3 ± 1.0 (27.4 – 33.9) | 0.53 |
| Platelet count (x 10<sup>3</sup>/μl) | 258 ± 12 (221 – 301) | 240 ± 13 (189 – 282) | 0.39 |

Values are expressed as mean ± standard error (range) or number (%). <sup>a</sup>ECMO, extracorporeal membrane oxygenation (ECMO). <sup>b</sup> BMI, body mass index. <sup>c</sup> IPF, idiopathic pulmonary fibrosis. <sup>d</sup> COPD, chronic obstructive pulmonary. <sup>e</sup> LAM, lymphangioleiomyomatosis. <sup>f</sup> CTD-ILD, connective tissue disease-associated interstitial lung disease. <sup>g</sup> FVC, forced vital capacity. <sup>h</sup> FEV1, forced expiratory volume in one second. <sup>i</sup> PT-INR, prothrombin time–international normalized ratio. <sup>j</sup> APTT, activated partial thromboplastin time.

### Operative characteristics

Table 2 shows the operative characteristics. There was no significant difference in the operative and cold ischemic time (CIT) between the 2 groups. The mean intraoperative ECMO time in the ECMO group was 487 ± 14 (443 – 557) minutes. The intraoperative blood loss in the ECMO group (1728 ± 734) tended to be greater than that in the control group (774 ± 308); however, the difference was not statistically significant (*p* = 0.36). In the control group, 1 patient (20.0%) intraoperatively required over 1,000 mL of blood transfusion of RBC. On the other hand, 3 patients (42.9%) were treated with over 1,000 mL of intraoperative blood transfusion of RBC in the ECMO group. The mean amount of transfusion of RBC in the ECMO group (920 ± 319) tended to be greater than that in the control group (336 ± 210); however, the difference was not statistically significant (*p* = 0.28). In the control group, 1 patient (20.0%) needed over 1,000 mL of fresh frozen plasma (FFP) transfusion. In the ECMO group, the requirement of FFP was over 1,000 mL in 2 patients (28.6%). No patient in the control group and 1 patient (14.3%) in the ECMO group required intraoperative transfusion of platelet. Table 2 shows the inflow and outflow cannula sizes in each case in the ECMO group. The cases with relatively smaller outflow cannula (case #5 and 12) tended to require more transfusion products.

**Table 2.** Operative characteristics.

| Case # | Group | OP <sup>a</sup> time (min) | CIT <sup>b</sup> (min) | Intra-OP ECMO time <sup>c</sup> (min) | Cannula size (Fr) |  | Intra-OP blood loss (mL) | Pleural adhesion grade <sup>f</sup> | Intra-OP blood transfusion |  |  |
| --- | --- | --- | --- | --- | --- | --- | --- | --- | --- | --- | --- |
|  |  |  |  |  | Inflow | Outflow |  |  | RBC <sup>e</sup> (mL) | FFP <sup>d</sup> (mL) | Platelet (mL) |
| 4 | Control | 365 | 361 | - | - | - | 249 | None | 0 | 0 | 0 |
| 6 | Control | 383 | 456 | - | - | - | 644 | Severe | 280 | 480 | 0 |
| 7 | Control | 366 | 457 | - | - | - | 307 | None | 0 | 0 | 0 |
| 8 | Control | 452 | 480 | - | - | - | 2122 | Severe | 1400 | 1200 | 0 |
| 9 | Control | 430 | 564 | - | - | - | 550 | Moderate | 280 | 0 | 0 |
| 1 | ECMO | 504 | 457 | 459 | 22 | 22 | 2332 | None | 1680 | 2880 | 0 |
| 2 | ECMO | 423 | 453 | 517 | 19.5 | 18 | 547 | Moderate | 280 | 0 | 0 |
| 3 | ECMO | 356 | 447 | 455 | 21 | 21 | 869 | None | 560 | 480 | 0 |
| 5 | ECMO | 482 | 656 | 557 | 21 | 15 | 6228 | Severe | 1960 | 1440 | 200 |
| 10 | ECMO | 380 | 457 | 443 | 21 | 16.5 | 748 | Mild | 0 | 0 | 0 |
| 11 | ECMO | 409 | 550 | 485 | 21 | 16.5 | 177 | None | 0 | 0 | 0 |
| 12 | ECMO | 468 | 539 | 492 | 16.5 | 15 | 1198 | None | 1960 | 480 | 0 |
|  |  | *P* = 0.29 | *P* = 0.34 |  |  |  | *P* = 0.36 |  | *P* = 0.28 | *P* = 0.44 |  |

<sup>a</sup> OP, operative. <sup>b</sup> CIT, cold ischemia time. <sup>c</sup> ECMO, extracorporeal membrane oxygenation (ECMO). <sup>d</sup> RBC, red blood cell. <sup>e</sup> FFP, fresh frozen plasma. <sup>f</sup> Pleural adhesion grade: severe, 50% to whole thoracic cavity; moderate, partial (only band adhesions) to < 50%; mild, no to partial adhesion; none, no adhesion.

### Intraoperative ADAMTS13 activity and vWF:Ag

Fig. 3A shows the comparisons of ADAMTS13 activity between the 2 groups in the intraoperative period (before lung transplantation, before reperfusion, and at the end of surgery). There was no significant difference in the ADAMTS13 activity between the ECMO and the control group in the intraoperative period. Fig. 3B shows the comparisons of vWF:Ag between the 2 groups in the intraoperative period. The level of vWF:Ag, representing a total amount of vWF multimers, was expressed as its ratio to that in the standard human plasma because the value of vWF:Ag varies significantly according to the age, ABO blood group, and so forth [18]. There was no significant difference in vWF:Ag between the two groups during the intraoperative period.

**Figure 3.**
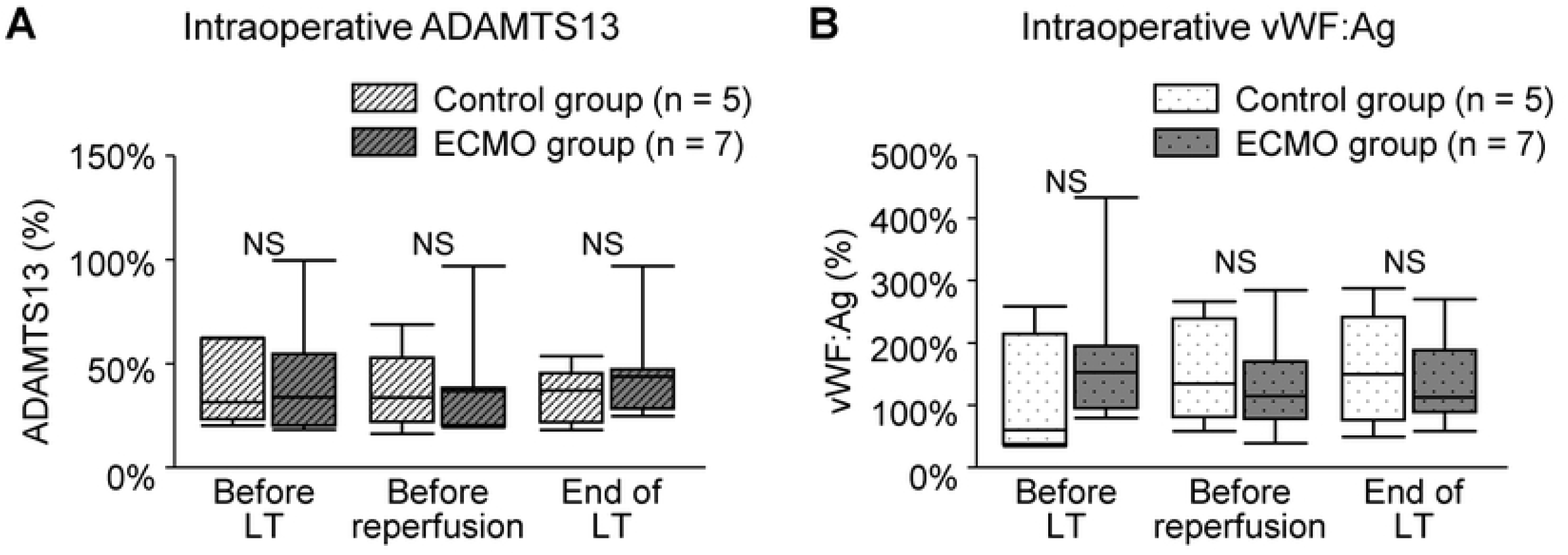
Intraoperative ADAMTS13 activity and von Willebrand factor antigen (vWF:Ag) For intraoperative analysis, blood samples were collected from the patients immidiately before lung transplantation, before reperfusion (immediately before reperfusion of the lung graft), and at the end of surgery. These time points are indicated as “Before LT,”” Before reperfusion,” and “End of LT,” respectively (LT, lung transplantation). The data are represented as a box and whiskers plot. White boxes represent the control group (n = 5, white boxes with stripes for ADAMTS13 activity, white boxes with dots for vWF:Ag), and gray boxes are the extracorporeal membrane oxygenation (ECMO) group (n = 7, gray boxes with stripes for ADAMTS13 activity, gray boxes with dots for vWF:Ag). The middle horizontal line represents the median. The upper and lower borders of the box represent the upper and lower quartiles. The upper and lower whiskers represent the maximum and minimum values. (A) Intraoperative ADAMTS13 activity. There was no significant difference in the ADAMTS13 activity between the control and ECMO group in the perioperative period. The mean ADAMTS13 activity before LT (control group vs. ECMO group), 41.6 ± 8.2% vs. 45.2 ± 10.1%, *p* > 0.05; before reperfusion, (control group vs. ECMO group), 37.6 ± 8.0% vs. 41.7 ± 9.3%, *p* > 0.05; at the end of LT, (control group vs. ECMO group), 35.3 ± 5.5% vs. 47.4 ± 8.5%, *p* > 0.05. (B) Intraoperative vWF:Ag. There was no significant difference in the vWF:Ag between the control and ECMO group in the intraoperative period. The mean vWF:Ag activity at before LT (control group vs. ECMO group), 114.4 ± 40.1% vs. 181.2 ± 42.8%, *p* > 0.05; before reperfusion, (control group vs. ECMO group), 157.5 ± 34.1% vs. 136.9 ± 27.9%, *p* > 0.05; at the end of LT, (control group vs. ECMO group), 158.9 ± 37.0% vs. 137.0 ± 25.4%, *p* > 0.05.

### Intraoperative vWF large multimer index

Fig. 4A shows the vWF large multimer index comparisons between the 2 groups in the intraoperative period. The vWF large multimer index in the control group maintained a level around 100% throughout the perioperative period (mean vWF large multimer index; before LT, 76.7 ± 10.0%; before reperfusion, 100.1 ± 10.9%; end of LT, 112.6 ± 10.6%). On the other hand, the vWF large multimer index in the ECMO group decreased before reperfusion and at the end of LT (mean vWF large multimer index; before LT, 93.8 ± 7.0%; before reperfusion, 74.7 ± 8.4%; end of LT, 75.8 ± 7.4%). The vWF large multimer index at the end of LT was significantly lower in the ECMO group than that in the control group (*p* < 0.05). There were 3 patients who did not receive intraoperative transfusion of FFP in the control group and the ECMO group, respectively.

**Figure 4.**
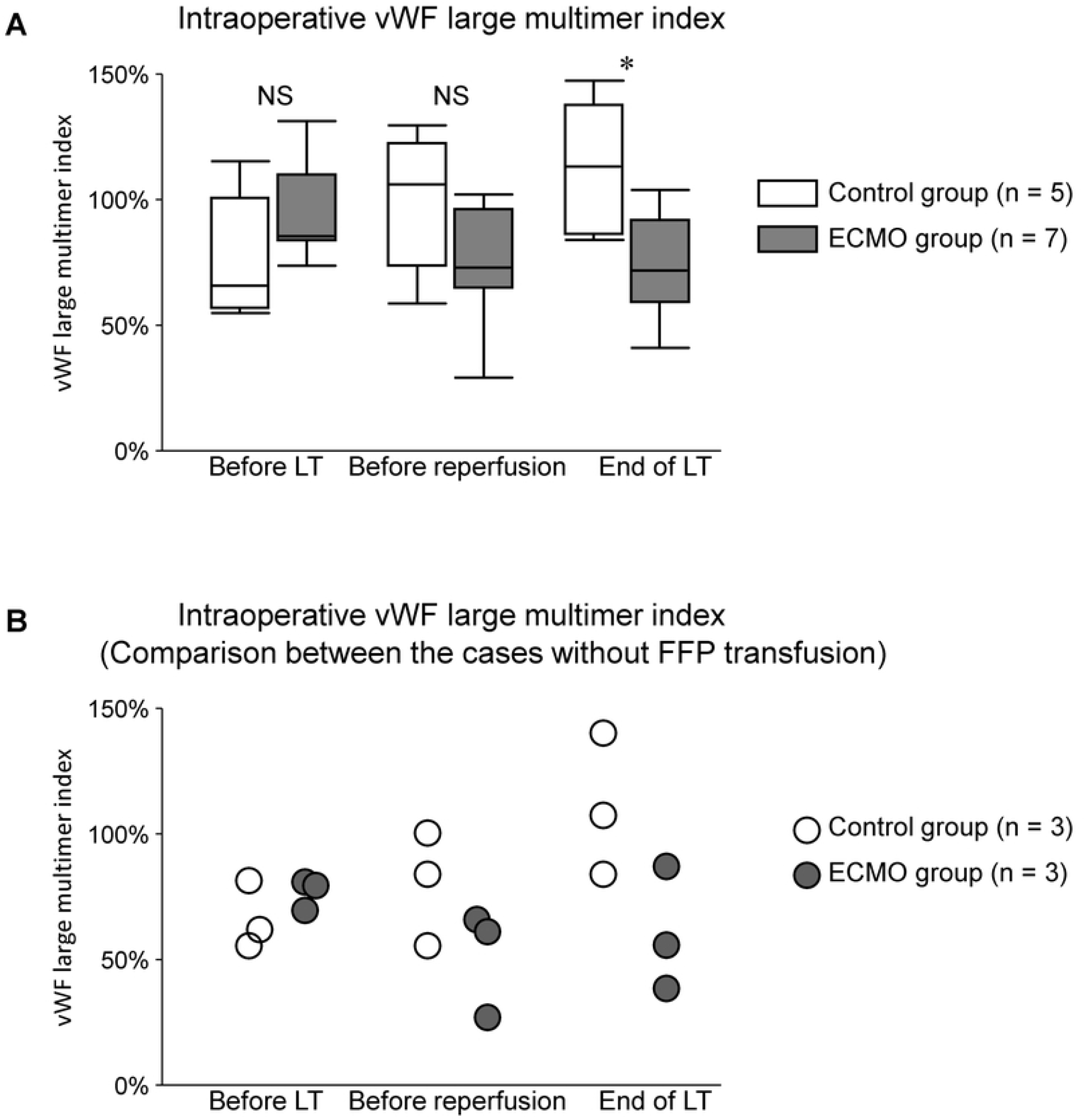
Intraoperative von Willebrand factor (vWF) large multimer index. For intraoperative analysis, blood samples were collected from the patients right before lung transplantation, before reperfusion (immediately before reperfusion of the lung graft), and at the end of surgery. These time points are indicated as “Before LT,”” Before reperfusion,” and “End of LT,” respectively (LT, lung transplantation). (A) Intraoperative vWF large multimer index. The data are represented as a box and whiskers plot. White boxes represent the control group (n = 5), and gray boxes are the extracorporeal membrane oxygenation (ECMO) group (n = 7). The middle horizontal line represents the median. The upper and lower borders of the box represent the upper and lower quartiles. The upper and lower whiskers represent the maximum and minimum values. The mean von Willebrand factor (vWF) large multimer index at before LT (control group vs. ECMO group), 76.7 ± 10.0% vs. 93.8 ± 7.0%, *p* > 0.05; at before reperfusion, (control group vs. ECMO group), 100.1 ± 10.9% vs. 74.7 ± 8.4%, *p* > 0.05; at end of LT, (control group vs. ECMO group), 112.6 ± 10.6% vs. 75.8 ± 7.4%, *p* < 0.05. (B) Intraoperative vWF large multimer index: comparison between the cases without fresh frozen plasma (FFP) transfusion. There were 3 patients in the control group and the ECMO group, respectively who did not receive intraoperative transfusion of FFP. The vWF large multimer index of each case in the control group was 66.8%/86.6%/59.5% before LT, 89.0%/106.0%/59.5% before reperfusion, and, 89.0%/113.5%/147.7% at the end of LT. The vWF large multimer index of each case in the ECMO group was 84.6%/85.9%/74.2% before LT, 70.6%/65.5%/29.9% before reperfusion, and 92.3%/59.8%/41.8% at the end of LT.

In order to exclude the influence of FFP transfusion on the vWF large multimer indices, we compared the indices in these patients. Fig. 4B shows each patient’s intraoperative vWF large multimer index without intraoperative FFP transfusion from the control and the ECMO group. The vWF large multimer index of each case in the control group was 66.8%/86.6%/59.5% at before LT, 89.0%/106.0%/59.5% before reperfusion, and, 89.0%/113.5%/147.7% at the end of LT. The vWF large multimer index of each case in the ECMO group was 84.6%/85.9%/74.2% before LT, 70.6%/65.5%/29.9% before reperfusion, and 92.3%/59.8%/41.8% at the end of LT. Namely, intraoperative vWF large multimer indices decreased to as low as 30–40% in the ECMO group without FFP transfusion whereas that in the control group was maintained at > 60% during SLT surgery.

### Postoperative characteristics and mortality

Table 3 shows each group’s postoperative 30-day, 90-day, and 1-year mortality. No patient was on ECMO at the intensive care unit upon arrival in the control group, and 2 patients (28.6%) were not weaned from VV ECMO in the ECMO group. There was no re-exploration thoracotomy for bleeding in either group. There was no mortality case in the control group within 1 year after transplantation. There was one 90-day mortality case in the ECMO group. The cause of the mortality was PGD.

**Table 3.** Postoperative characteristics and mortality.

|  | Control group<br>(*n* = 5) | ECMO <sup>a</sup> group<br>(*n* = 7) |
| --- | --- | --- |
| ECMO on ICU <sup>b</sup> arrival | 0 (0%) | 2 (28.6%) |
| Re-exploration thoracotomy for bleeding | 0 (0%) | 0 (0%) |
| Mortality |  |  |
| 30-day | 0 (0%) | 0 (0%) |
| 90-day | 0 (0%) | 1 (14.3%) <sup>b</sup> |
| 1-year | 0 (0%) | 0 (0%) |

Values are expressed as numbers (%). <sup>a</sup> ECMO, extracorporeal membrane oxygenation. <sup>b</sup> ICU, intensive care unit. <sup>c</sup> The cause of death was primary graft dysfunction.

## Discussion

This is the first study that quantitatively analyzed the intraoperative changes in vWF large multimers in patients who underwent lung transplantation. We demonstrated that vWF large multimer index, which is regarded as the most reliable predictor of AvWS in recent years, was lower in the patients who underwent VV ECMO support during SLT compared to those without ECMO at the end of the surgery. However, the vWF large multimer index was maintained at > 75% in average at the end of SLT in the ECMO group, which did not significantly affect the intra- and postoperative outcomes including blood loss, blood transfusion, and re-exploration thoracotomy for bleeding.

In the present study, the ADAMTS13 activity and vWF:Ag showed no significant change by intraoperative VV ECMO use. On the other hand, there was a significant decrease in the vWF large multimer index at the end of the surgery in SLT. However, the mean vWF large multimer index at the end of surgery was relatively maintained at 75.8% in the ECMO group. The intraoperative blood loss and the required amounts of intraoperative transfusion products in the ECMO group tended to be greater than those in the control group, but the difference was not statistically significant. We previously reported a more severe decrease in the vWF large multimer index in 5 patients with acute cardiovascular diseases treated with VA ECMO [10].

These patients exhibited vWF large multimer indices of 20.8% at 24 hours, 51.0% at 23 hours, 27.6% at 47 hours, 28.8% at 19 hours, and 31.0% at 82 hours after VA ECMO initiation [10], and 2 of the 5 patients had massive bleeding events, while the oozing type of bleeding at the skin of the sites of cannula insertion was observed in all 5 patients, indicating an apparent hemostatic disorder [10].

The results regarding AvWS in our study were also quite different from those in a previous study reported by Kalbhenn et al. They demonstrated that patients receiving VV ECMO support for respiratory failure were all diagnosed with AvWS, as shown by the ratio of the collagen-binding capacity (vWF:CB) to vWF:Ag decreasing to about 0.25 in average (vWF:CB/ vWF:Ag, normal: > 0.7). They also reported that 17 of the 18 patients developed bleeding complications from 3 to 30 days after ECMO[14]. The different results in our study from those in previous studies may be primarily explained by the short duration of time of ECMO use in our study. The relatively low threshold for the use of intraoperative FFP transfusion in our cohort, the thicker venous outflow cannulas in VV ECMO compared to those in VA ECMO in general [10], and the lower blood flow rate of 1.5 – 2.5 L/min in the present study compared with that in VA ECMO used for patients with severe respiratory failure may also be responsible for the difference.

We acknowledge that there are several limitations in the present study. First, it was a single-center analysis with a small number of patients, and thus we cannot accurately compare the outcome of each group. Second, there were a few patients receiving an intraoperative transfusion of FFP in both groups. Therefore, an accurate comparison of the vWF large multimer indices was impossible. Whereas we compared the vWF large multimer indices in the patients without intraoperative FFP transfusion between the control and the ECMO group, the number of patients was not enough for statistical analysis. Third, we were not able to enroll a large enough number of patients to undergo lung transplantation under CPB or VA ECMO in this study, which would be a future challenges.

In conclusion, intraoperative use of VV ECMO for SLT caused a decrease in the vWF large multimer index. However, the vWF large multimer index in the present study remained at an average of > 75% and was as low as 30–40% at the end of surgery with VV ECMO, and its impact on bleeding complications was limited.

## Data Availability

All relevant data are within the manuscript and its Supporting Information files.

## Acknowledgments

This research was supported by Japan Agency for Medical Research and Development under Grant Number 20ek0109370h0003. A part of this study was conducted as collaborative research with Sysmex Corporation (Kobe, Japan).

## Notes

### Competing Interest Statement

The authors have declared no competing interest.

### Clinical Trial

UMIN Clinical Trials Registry (UMIN-CTR), UMIN000018135

### Clinical Protocols

https://center6.umin.ac.jp/cgi-open-bin/ctr_e/ctr_view.cgi?recptno=R000019166

### Funding Statement

HH, Japan Agency for Medical Research and Development (Grant Number 20ek0109370h0003)

### Author Declarations

The Ethics Committee of Tohoku University Graduate School of Medicine

